# A Rapid Review and Meta-Analysis of the Asymptomatic Proportion of PCR-Confirmed SARS-CoV-2 Infections in Community Settings

**DOI:** 10.1101/2020.05.20.20108183

**Authors:** Sarah Beale, Andrew Hayward, Laura Shallcross, Robert W Aldridge, Ellen Fragaszy

**Affiliations:** UCL Institute of Health Informatics, 222 Euston Rd, London NW1 2DA; UCL Research Department of Epidemiology & Public Health, 1-19 Torrington Place, London WC1E 7HB; LSHTM Department of Infectious Disease Epidemiology, Keppel Street, London WC1E 7HT

## Abstract

**Background:** Up to 80% of active SARS-CoV-2 infections are proposed to be asymptomatic based on cross-sectional studies. However, accurate estimates of the asymptomatic proportion require systematic detection and follow-up to differentiate between truly asymptomatic and pre-symptomatic cases. We conducted a rapid review and meta-analysis of the asymptomatic proportion of PCR-confirmed SARS-CoV-2 infections based on methodologically-appropriate studies in community settings.

**Methods:** We searched Medline and EMBASE for peer-reviewed articles, and BioRxiv and MedRxiv for pre-prints published before 25/08/2020. We included studies based in community settings that involved systematic PCR testing on participants and follow-up symptom monitoring regardless of symptom status. We extracted data on study characteristics, frequencies of PCR-confirmed infections by symptom status, and (if available) cycle threshold/genome copy number values and/or duration of viral shedding by symptom status, and age of asymptomatic versus (pre)symptomatic cases. We computed estimates of the asymptomatic proportion and 95% confidence intervals for each study and overall using random effect meta-analysis.

**Findings:** We screened 1138 studies and included 21. The pooled asymptomatic proportion of SARS-CoV-2 infections was 23% (95% CI 16%-30%). When stratified by testing context, the asymptomatic proportion ranged from 6% (95% CI 0-17%) for household contacts to 47% (95% CI 21-75%) for non-outbreak point prevalence surveys with follow-up symptom monitoring. Estimates of viral load and duration of viral shedding appeared to be similar for asymptomatic and symptomatic cases based on available data, though detailed reporting of viral load and natural history of viral shedding by symptom status were limited. Evidence into the relationship between age and symptom status was inconclusive.

**Conclusion:** Asymptomatic viral shedding comprises a substantial minority of SARS-CoV-2 infections when estimated using methodologically-appropriate studies. Further investigation into variation in the asymptomatic proportion by testing context, the degree and duration of infectiousness for asymptomatic infections, and demographic predictors of symptom status are warranted.

## Background

Reports of asymptomatic SARS-CoV-2 infection and potential transmission^1,2,3^ have generated concern regarding the implications of undetected asymptomatic transmission on the effectiveness of public health interventions in the current COVID-19 pandemic^4^. However, estimating the proportion of asymptomatic SARS-CoV-2 infections with viral shedding is challenging as the majority of testing is carried out on symptomatic individuals^5^. Furthermore, longitudinal designs that include symptom follow-up are required to differentiate truly asymptomatic cases, i.e. those that never develop symptoms during infection, from pre-symptomatic cases, i.e. those that shed virus and therefore test positive prior to symptom onset (see Figure 1). While asymptomatic viral shedding has been suggested to comprise up to ∼80% of SARS-CoV-2 infections ^6,7,8^, data informing these figures are largely confined to cross-sectional reports that cannot distinguish truly asymptomatic cases from those who are pre-symptomatic at the point of testing (see Figure 1). Interchangeable use of these concepts, i.e. asymptomatic and pre-symptomatic, precludes accurate estimation of the asymptomatic proportion of potentially infectious SARS-CoV-2 cases. Detectible SARS-CoV-2 shedding based on reverse transcriptase polymerase chain reaction (PCR) testing cannot conclusively establish infectiousness in the absence of viral culture ^9,10^. However, PCR cycle threshold values provide an informative estimate of viral load and, by extension, probable infectiousness ^9^; consequently, PCR-confirmed infection can provide a useful and accessible indicator of potentially infectious cases, including those without symptoms, for epidemiological modelling.

**Figure 1.**
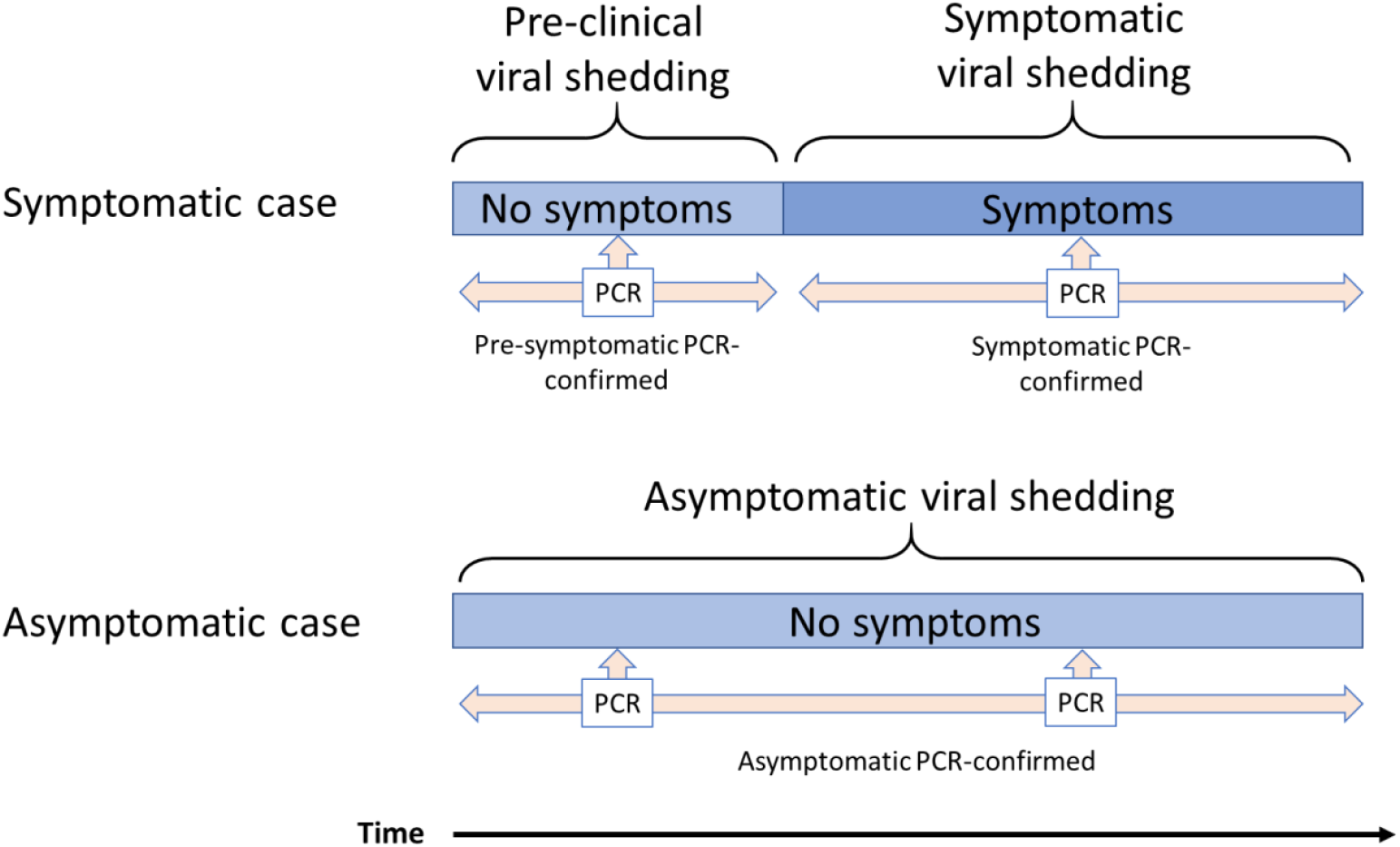
Timeline of Symptom Development and Viral Shedding in Relation to Timing of Virological Testing. ***Note:*** This figure demonstrates two trajectories of symptom development in cases with detectable viral shedding. The symptomatic case trajectory comprises a period of pre-clinical viral shedding, in which the individual demonstrates no symptoms but tests PCR positive (pre-symptomatic PCR-confirmed). These individuals subsequently develop symptoms and continue to shed virus (symptomatic PCR-confirmed). Consequently, cases with a symptomatic trajectory may appear to be asymptomatic if tested in the pre-clinical shedding period and not followed-up. Asymptomatic cases with viral shedding, conversely, test PCR positive and never go on to develop symptoms across the course of infection (asymptomatic PCR-confirmed).

Differences in demographic characteristics of asymptomatic versus symptomatic individuals are also poorly understood. Age is an important risk factor for COVID-19 severity, with greater risk of poor prognostic outcomes including mortality in older adults^11,12^. Consequently, asymptomatic infection may be less common with increasing age. Understanding the relationship between age and symptom status has important implications for public health interventions.

Given the widespread discussion and potential implications of asymptomatic transmission of SARS-CoV-2, we aimed to rapidly synthesize studies to estimate the asymptomatic proportion of PCR-confirmed cases in community settings (primary outcome). We also aimed to synthesize available data from these studies regarding viral load and duration of viral shedding in asymptomatic community cases compared to pre-symptomatic cases or those symptomatic from baseline (secondary outcome), and the relationship between symptom status and age (secondary outcome). We limited the review to include studies from community settings rather than hospitals and other medical facilities to prevent selection bias towards symptomatic cases. Only studies reporting PCR-confirmed cases rather than exclusive serological studies were included to estimate the proportion of asymptomatic SARS-CoV-2 infection with viral shedding. The review was not extended to estimate the overall asymptomatic proportion including non-shedding serological cases due to the limited number of serological studies, varying interpretation, and ongoing development of valid serological assays for SARS-CoV-2.

## Methodology

This review was reported in line with the PRISMA guidelines^13^. A protocol was not registered due to its status as a rapid review.

### Search Strategy

We used Ovid to search the Medline and EMBASE databases of peer-reviewed literature (2019-May 05 2020 and search repeated to include period of May 06 2020 to June 10 2020, and subsequently to include June 11 2020 to August 25 2020) using the following search terms for titles and abstracts: *(Coronavirus* OR Covid-19 OR SARS-CoV-2 OR nCoV) AND (asymptomatic)* AND (*polymerase chain reaction OR PCR OR laboratory-confirmed OR confirmed*). We also searched BioRxiv and MedRxiv for titles and abstracts of pre-print manuscripts using the terms *“Covid-19” + “asymptomatic”*. We hand-searched the reference lists of all included studies to identify any additional relevant literature.

### Selection Criteria

We included studies that met all of the following criteria: 1) human study; AND 2) presented original research or public health COVID-19 surveillance data; AND 3) available in English; AND 4) presented data on polymerase chain reaction (PCR) confirmed COVID-19 cases; AND 5) presented data on PCR testing of exposed or potentially exposed individuals regardless of symptom status (to avoid bias towards symptomatic cases); AND 6) had systematic follow-up at ≥ 1 time-point and reporting of symptom status among PCR confirmed cases (to differentiate pre-clinical shedding from truly asymptomatic cases); AND 7) presented data from a community setting (i.e. community and home contact tracing, population screening, traveller screening, community institutional settings such as care homes, schools, or workplaces, occupational exposure including healthcare workers). Studies were excluded if they met any of the following criteria: 1) studies or case series with <5 positive cases and/or <20 total cases (small sample size) due to likely low generalisability of asymptomatic proportions; OR 2) not possible to consistently ascertain the symptomatic status of participants across follow-up; OR 3) inadequate detail about testing strategy (i.e. not possible to discern if all cases were tested systematically); OR 4) recruitment/reporting of patients from acute healthcare settings (e.g. hospitals, medical facilities) due to selection bias towards symptomatic cases.

### Data Extraction and Analysis

One researcher performed the search and deduplication using Ovid, screened and selected studies, and extracted study details. Two researchers extracted primary outcome data independently and resolved any disagreement by consensus. We extracted the following variables of interest to assess the primary and secondary outcomes and the characteristics and quality of included studies: author names, year of publication, publication type (peer-reviewed article or pre-print), study design, study setting, study country of location, participant age (mean, median, or range as available), participant sex distribution, symptoms comprising symptomatic case definition, duration of symptom history at PCR-confirmation, duration of follow-up symptom monitoring, testing criteria, sample size, number of participants who underwent PCR testing, number of PCR-confirmed cases, number of confirmed cases who remained asymptomatic throughout follow-up, and cycle threshold or genome copy number values, viral culture results, duration of viral shedding for asymptomatic and pre-/symptomatic cases, and any available data regarding age or age distribution of asymptomatic versus (pre)symptomatic cases if reported.

We performed random-effects meta-analysis using the *metaprop* programme^14^ in Stata Version 15 to compute the study-specific and pooled asymptomatic proportion - the primary outcome of this review - with its 95% confidence intervals (Wilson score method) and 95% prediction intervals ^15^, applying the Freeman-Tukey transformation. We decided a-priori to use a random effects model to address heterogeneity. The asymptomatic proportion is given as the number of consistently asymptomatic confirmed cases divided by the total number of PCR-confirmed cases who received follow-up (Figure 2). It is important to note that the term asymptomatic proportion is sometimes used to alternatively refer to the asymptomatic proportion of all infections including those that do not shed virus and would not be PCR-confirmed (see Figure 2). To account for potential exposure-driven heterogeneity in asymptomatic proportion, we present findings stratified by testing context as well as overall. Testing context was subdivided into studies comprising exclusive household contacts of an index case, studies comprising contacts from other settings or those (potentially) exposed to an outbreak (including travellers returning from high-prevalence regions), and point prevalence surveys not specifically linked to an outbreak that had follow-up symptom monitoring.

**Figure 2.**
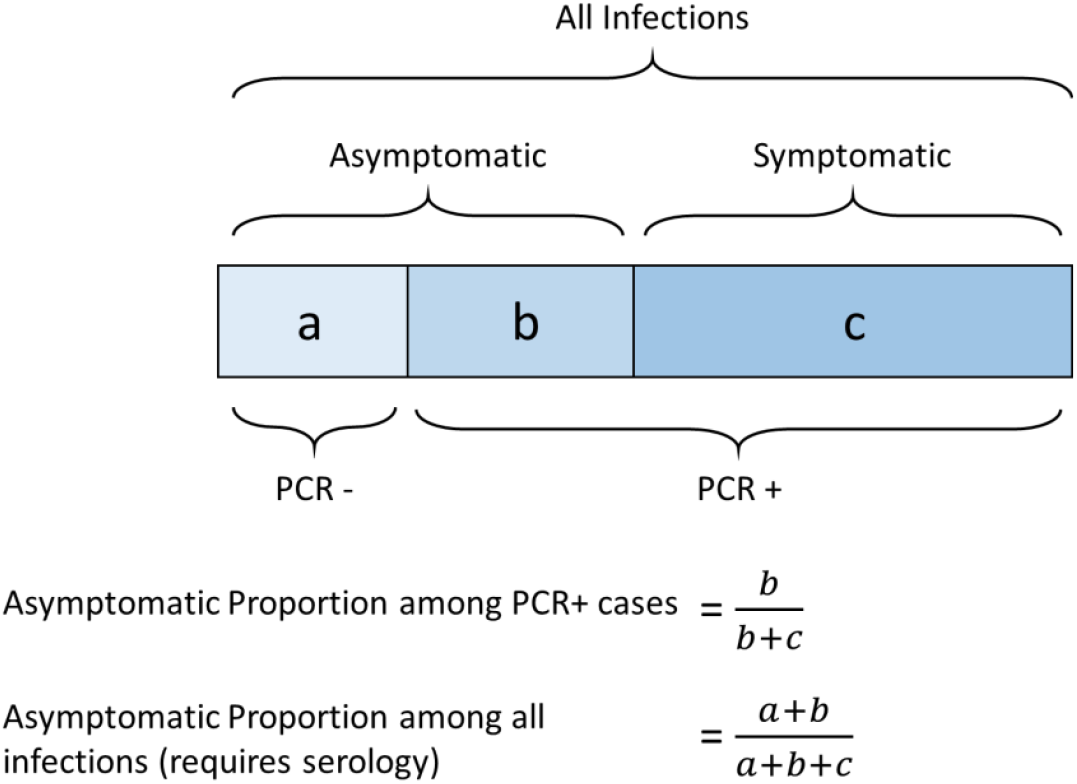
Summary Classification of Clinical and PCR Outcomes and Calculation of Asymptomatic Proportions.

We reported available findings regarding the viral load, duration of viral shedding, and age of asymptomatic and (pre)symptomatic cases, but did not conduct meta-analysis due to sparse reporting and inconsistencies in data presented.

### Risk of Bias Assessment

We assessed risk of bias based using criteria relevant to the topic of this review adapted from the Joanna Briggs Institute critical appraisal tool for prevalence studies^16^ (Table 1). Two researchers independently assessed the risk of bias for each included study and resolved any disagreement by consensus. Bias was assessed according to criteria described in Table 1, with studies graded as very low risk of bias if they were unlikely to have been affected by bias on any of the criteria, low if one criterion may have been affected, moderate if two may have been affected, and high if all three may have been affected. Risk of publication bias across included studies was assessed using a funnel plot and Egger’s test.

**Table 1.**
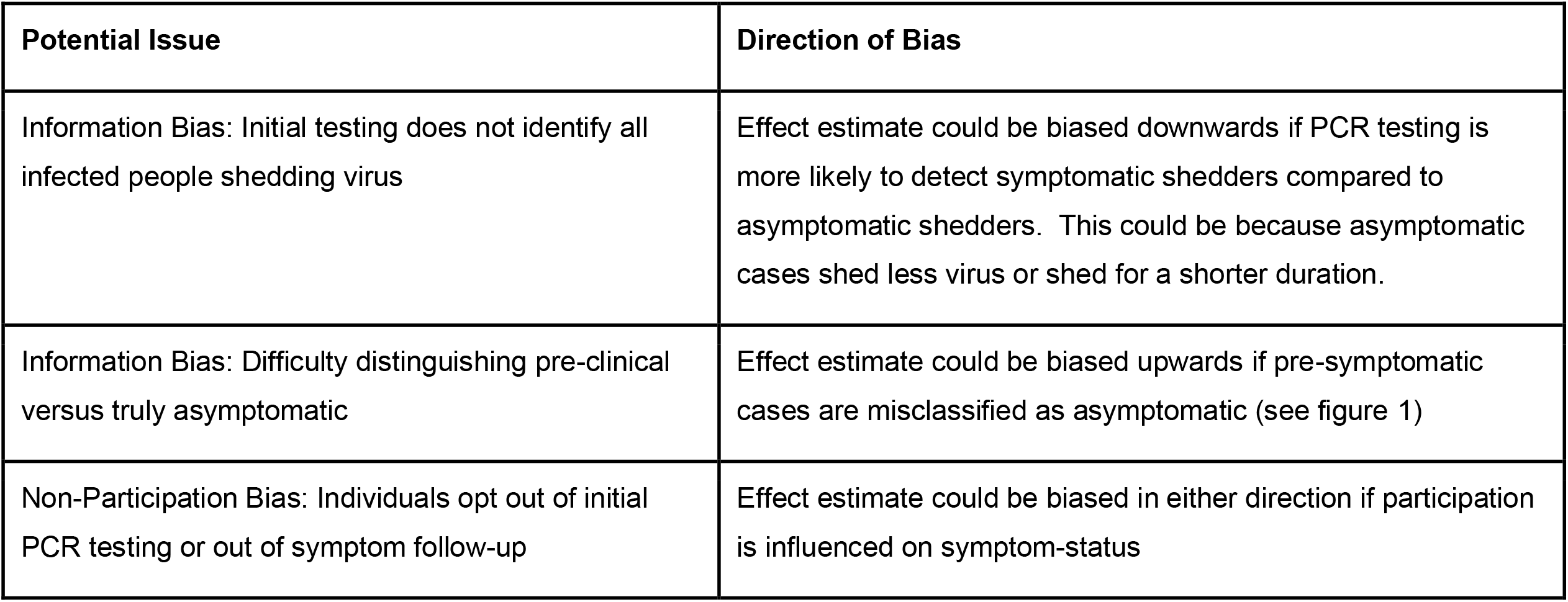
Risk of Bias Assessment.

## Results

### Records Identified

Figure 3 presents an adapted PRISMA flow diagram^13^ of the study selection procedure. The search yielded 1077 published articles indexed on OVID and 473 pre-prints. Following deduplication, we screened the titles and abstracts of 1138 published articles and pre-prints, of which we assessed the 133 full texts – including a relevant text identified through hand-search of the literature – and included 21 in the present review ^17-37^. Three of the 21 included studies comprised household contacts of confirmed cases^26,28,29^. A further 3 included studies were point prevalence surveys with symptom monitoring followup^34,35,36^, one of which was conducted in a general population sample^36^ and the remaining two in nursing home samples^34,35^. The remaining 15 studies involved participants with other epidemiological links to confirmed or suspected cases/outbreaks^17,18,19,20,21,22, 23,24,25,27,30,31,32,33^, including 5 studies based in nursing homes ^18,19,24,32,37^, and 1 study of healthcare workers with occupational exposure to confirmed cases^30^. The healthcare worker study was included as it comprised whole-facility testing following occupational exposure in healthcare workers rather than patients presenting to healthcare settings due to symptoms (see inclusion criteria). Studies were conducted across the following range of countries in Asia, Europe, and North America: China^22,25,26,27,29^, USA^18,19,28,32,36^, UK^24,35,37^, South Korea^17^, France^20^, Vietnam^21^, Brunei^23^, Italy^30^, Japan^31^, Hong Kong^33^, and Ireland^34^. Risk of bias was rated as very low for 2 studies ^30,33^, low for 15 studies^17-20,22,23,25-29,32,34-36^, and moderate for 4 studies^21,24,31,37^.

**Figure 3.**
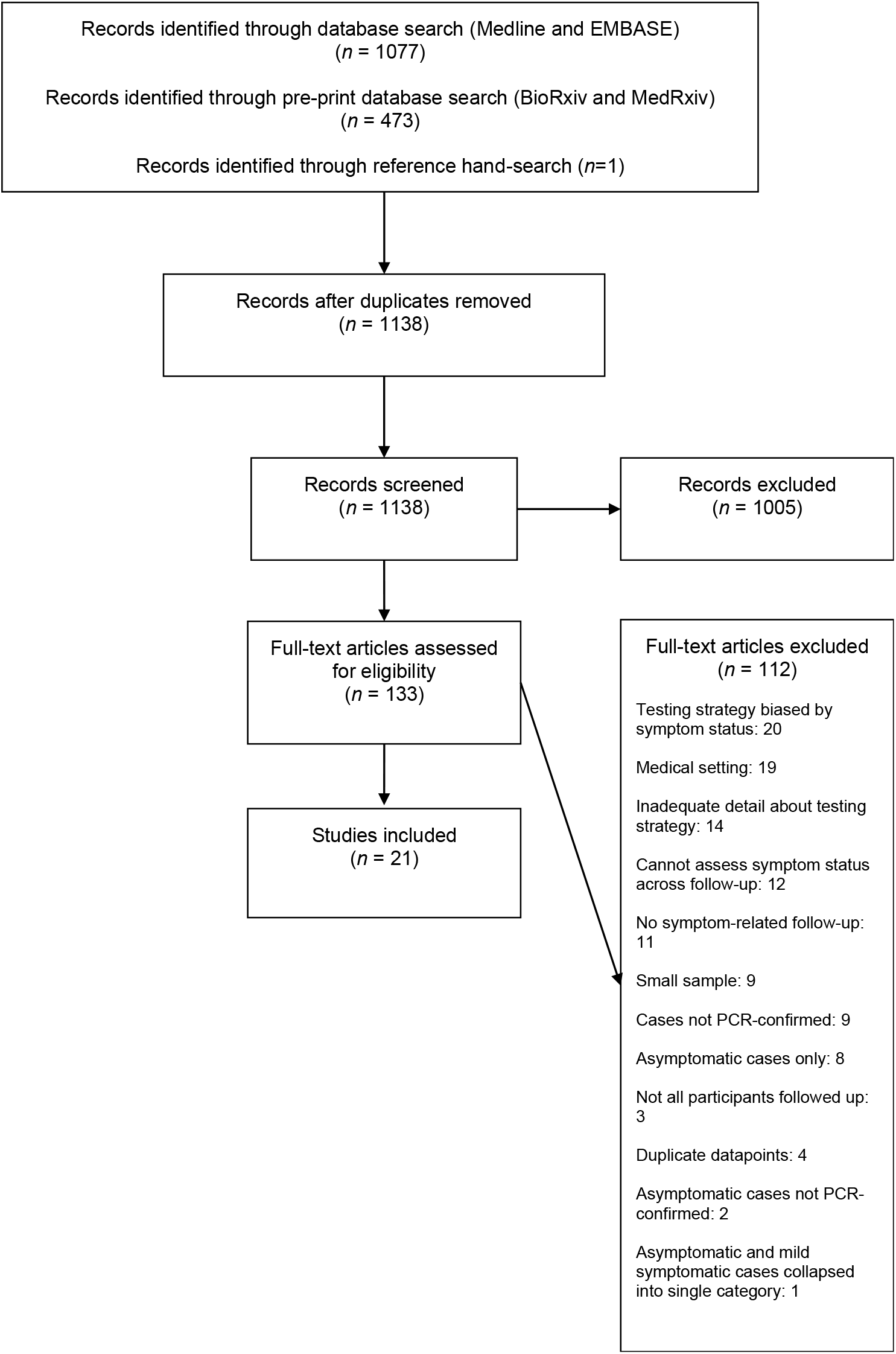
Adapted PRISMA Flow Diagram of Study Selection.

### Asymptomatic Proportion of PCR-Confirmed SARS-CoV-2 Infections in Community Settings

Estimates of the asymptomatic proportion of PCR-positive SARS-CoV-2 infections for included studies ranged from 0% (95% CI 0-0.8%; Yousaf et al., 2020^28^) to 91% (95% CI 73%-98%; Starling et al., 2020^35^). Table 2 reports all asymptomatic proportions with 95% confidence intervals for as well as details of included studies. Based on random-effects meta-analysis (Figure 4), the pooled estimate for the asymptomatic proportion was 23% (95% CI 16%-30%; 95% prediction interval 0.01-57%). There was high heterogeneity: *Q*(20)= 253.06, *p*<.001, τ^2^= 0.11, I^2^= 92.10%. Heterogeneity appeared to be partly influenced by testing context (test for subgroup heterogeneity: *Q*(2) = 10.49, *p*=0.01), but remained substantial within these subgroups. Household contact studies demonstrated the lowest asymptomatic proportion estimate of 6% (95% CI 0-17%; heterogeneity *Q*(2)= 12.09, *p*<.001 τ^2^= 0.07, I^2^= 83.46%), rising to 23% (95% CI 14-32%; *Q*(14)= 139.86, *p*<.001 τ^2^= 0.12, I^2^= 89.99%) for studies comprising participants with other epidemiological links to SARS-CoV-2 cases or outbreaks, and 47% (95% CI 21-75%; *Q*(2)= 47.16, *p*<.001 τ^2^= 0.23, I^2^= 95.76%) for point prevalence surveys with symptom follow-up and without direct links to outbreaks/cases. Data were limited for studies exclusively involving household contacts or point prevalence surveys (both *n*=3 studies).

**Table 2.**
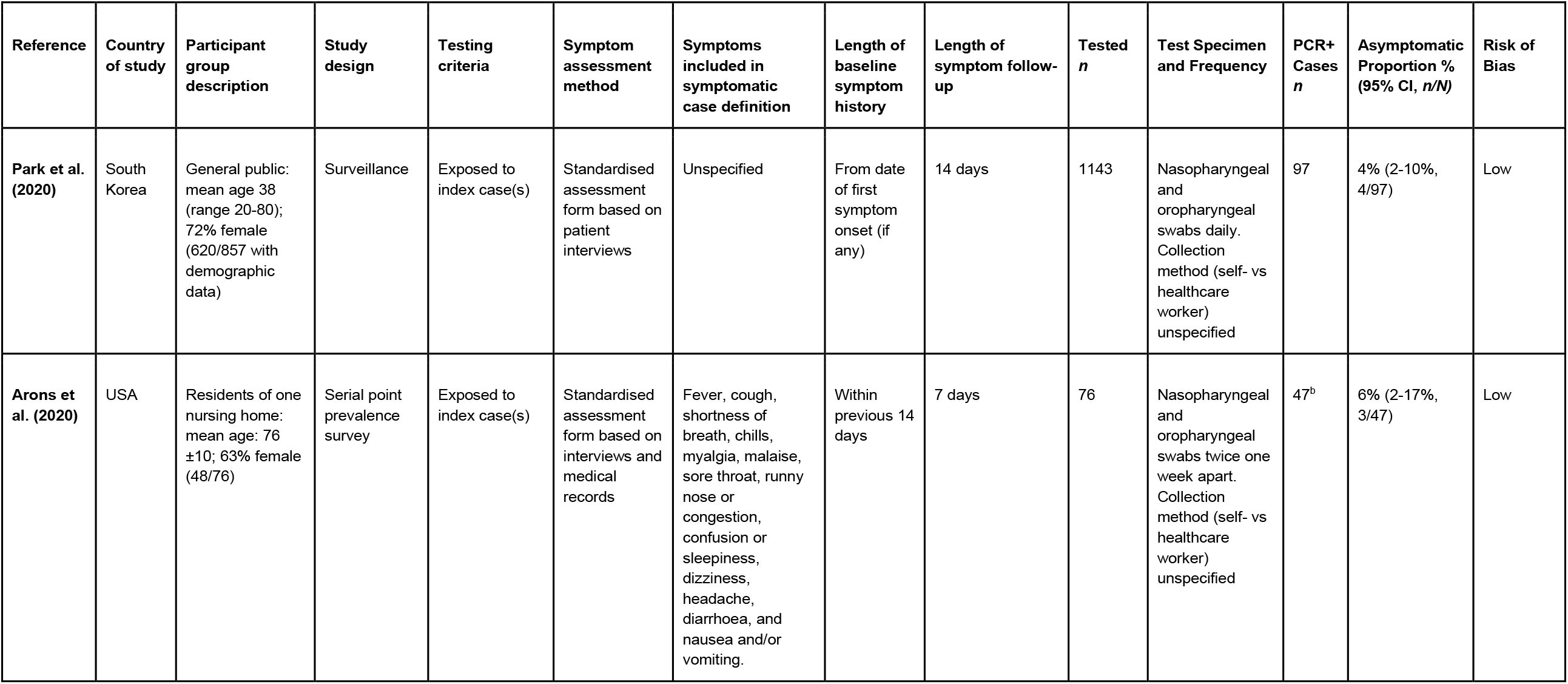
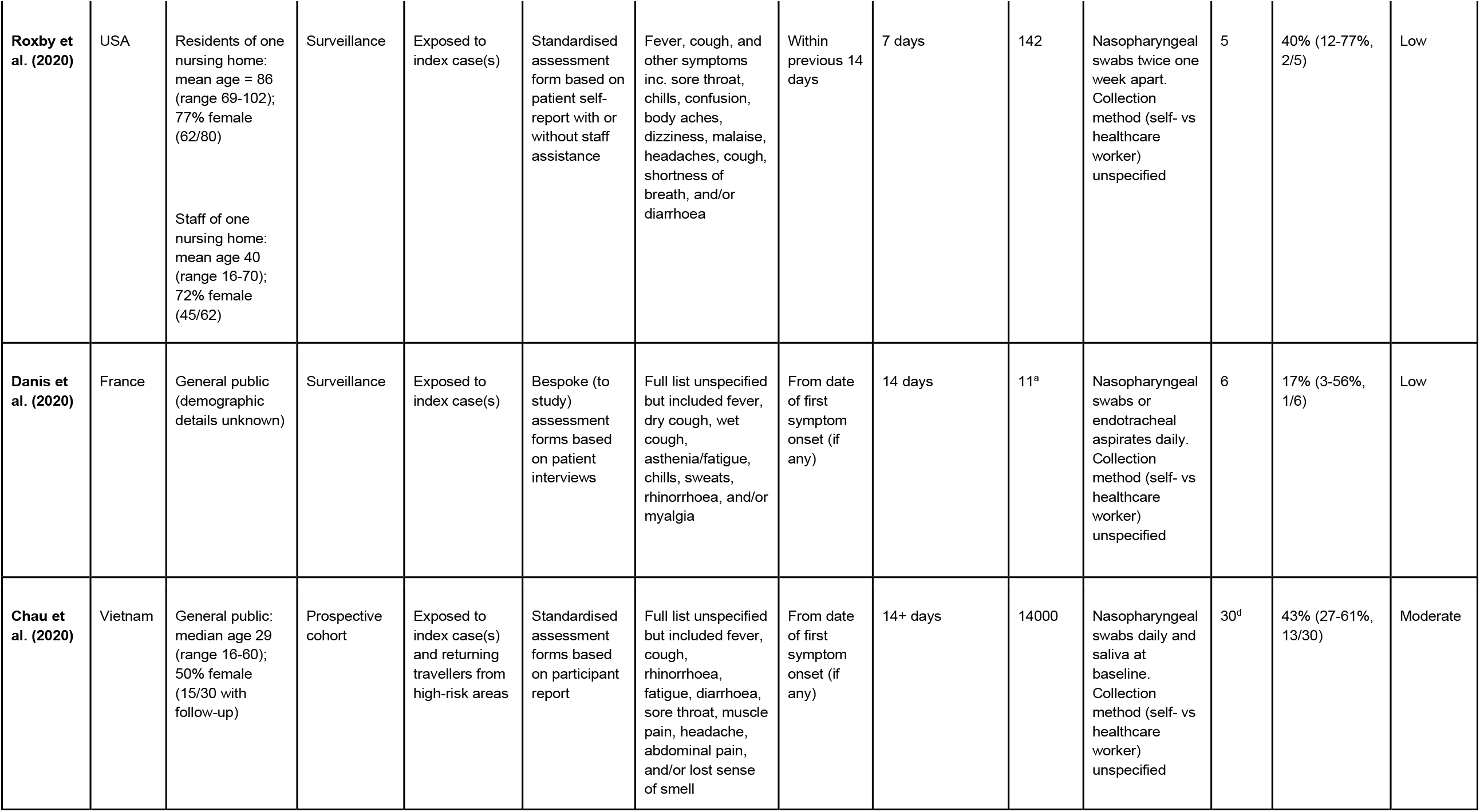
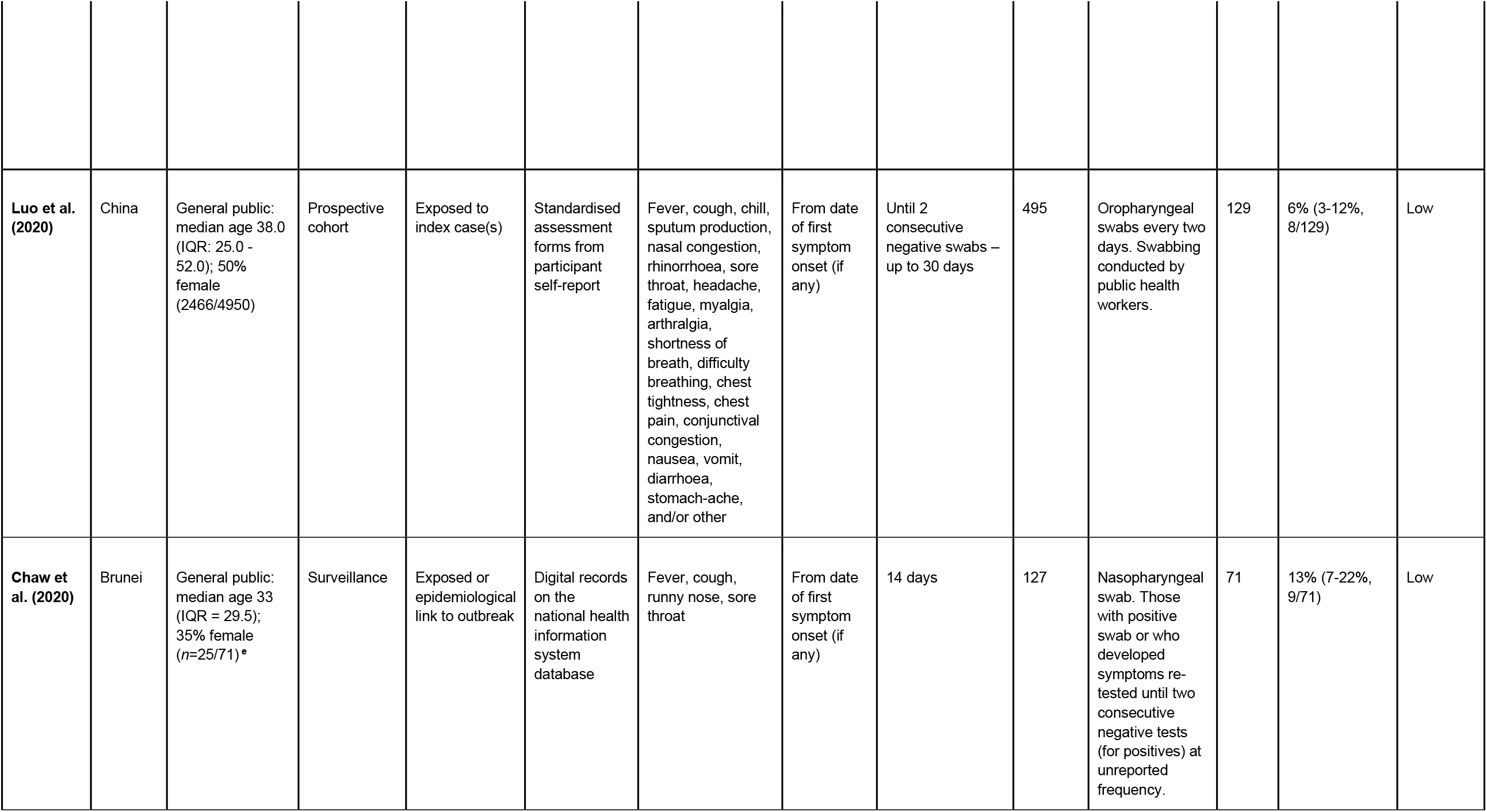
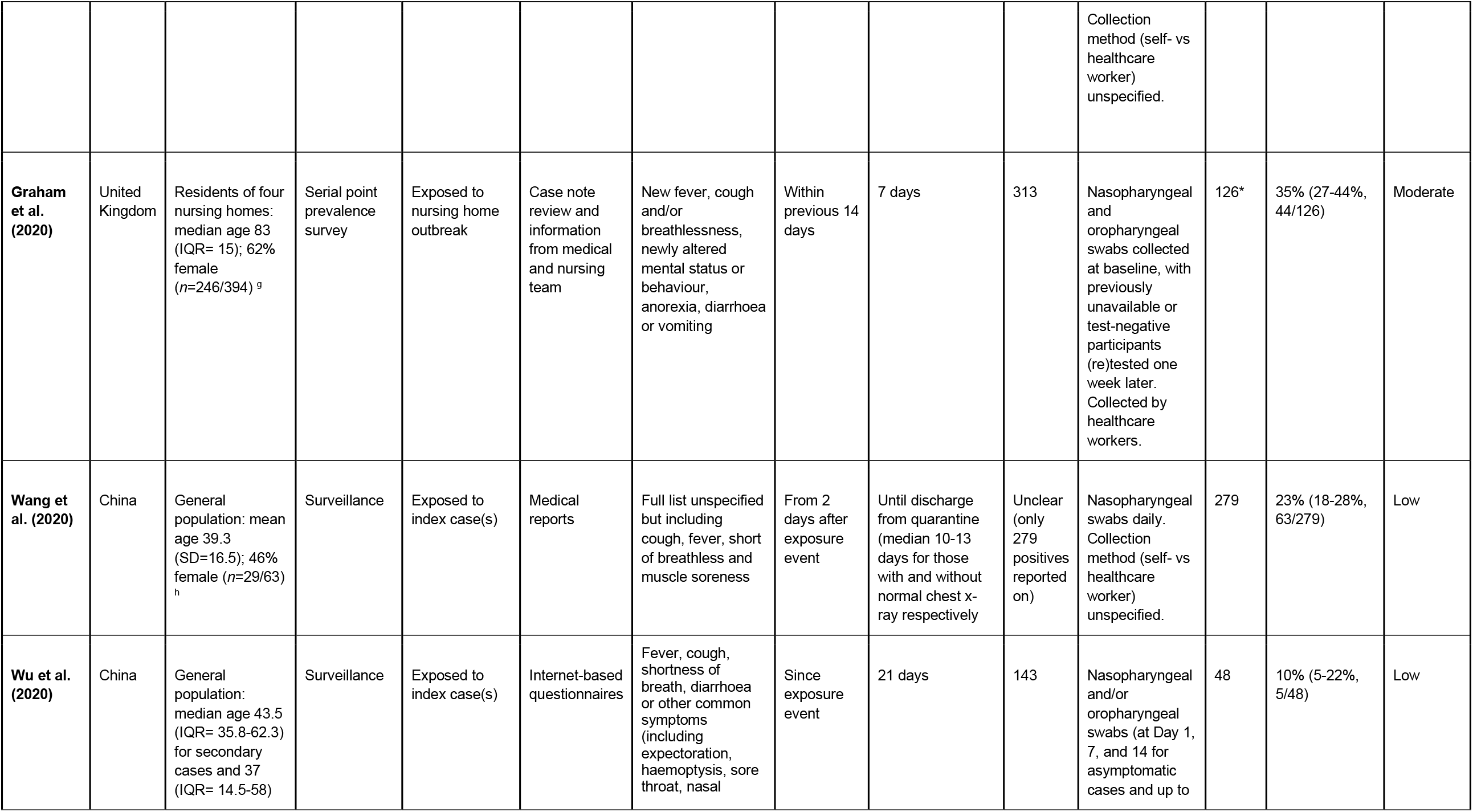
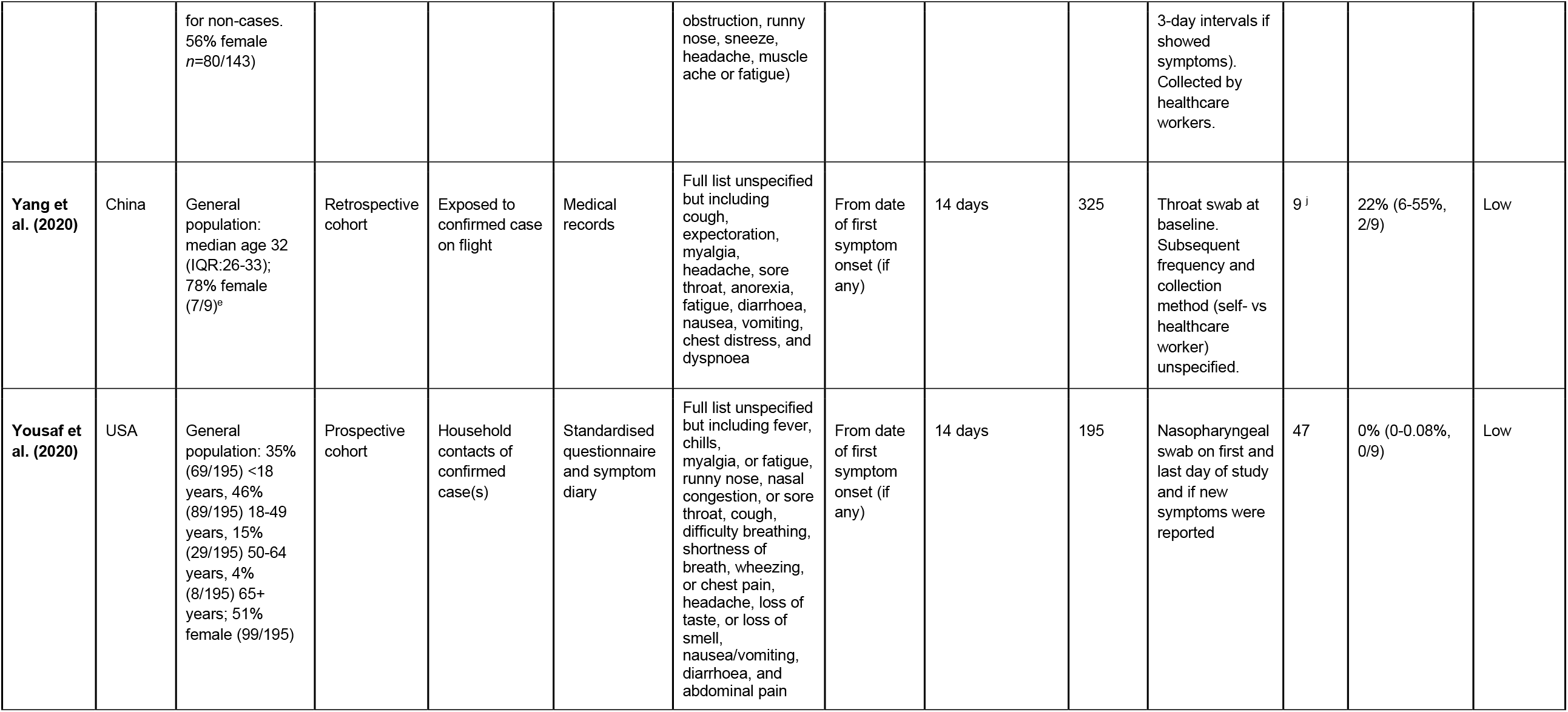
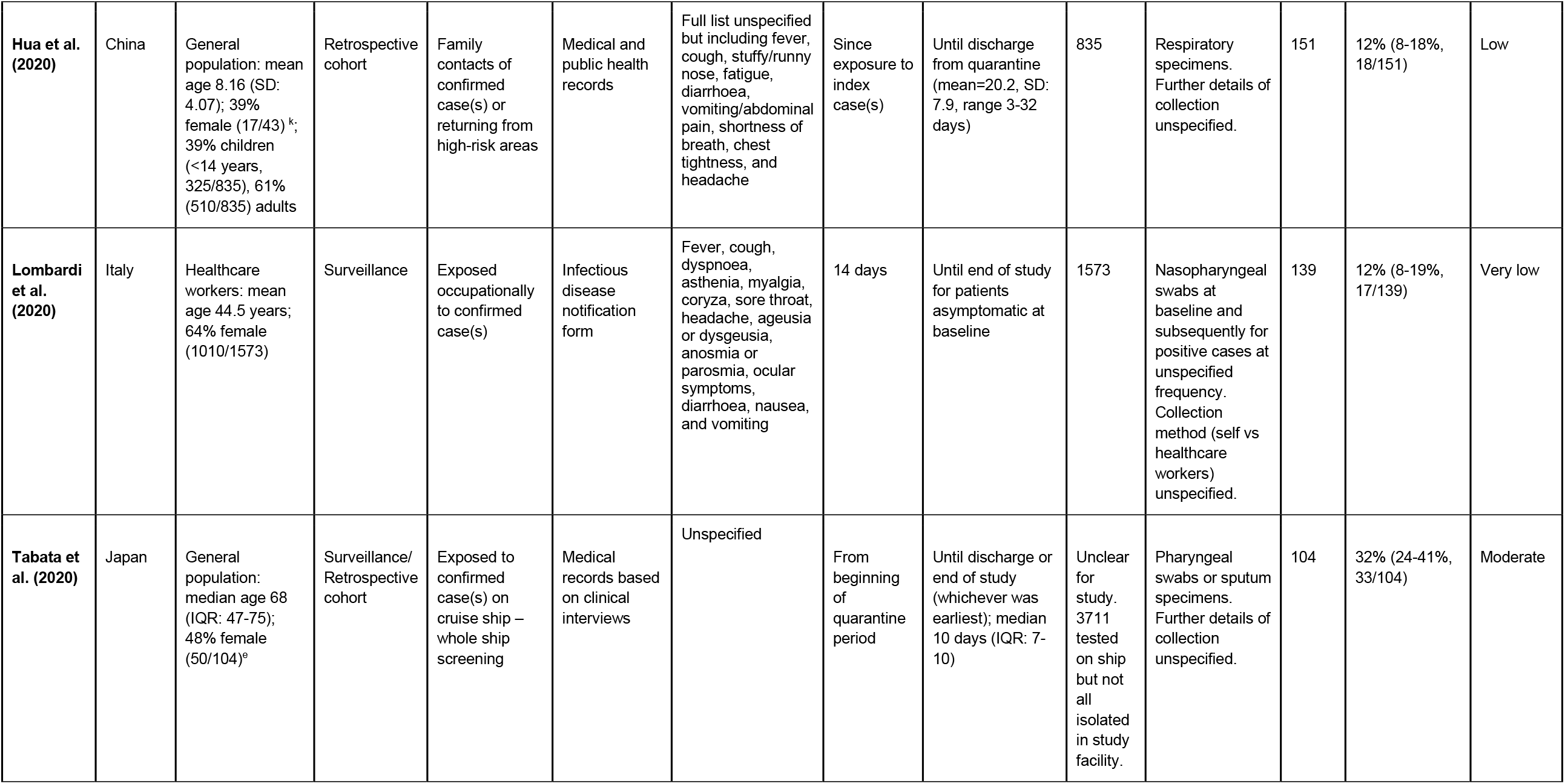
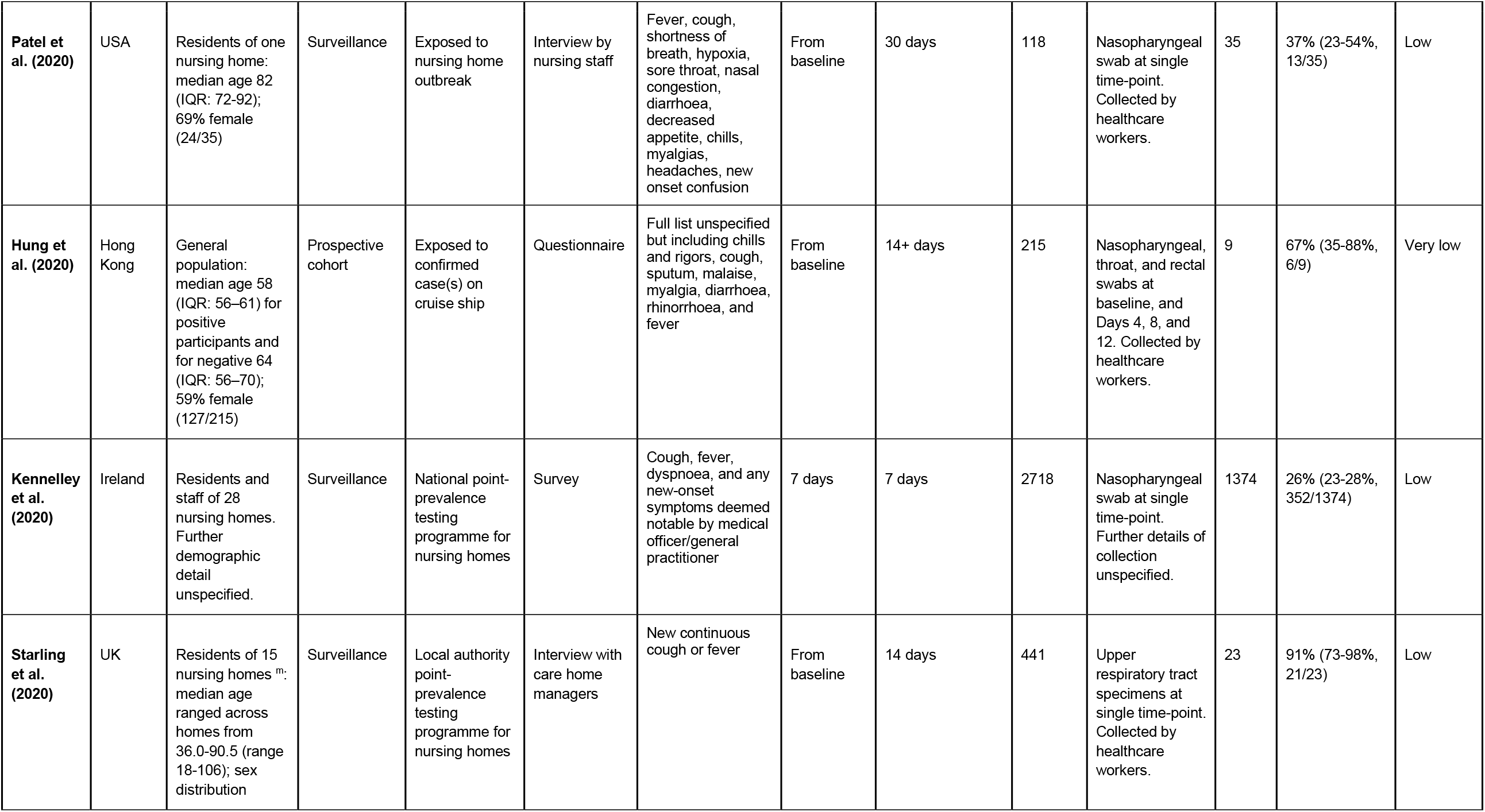
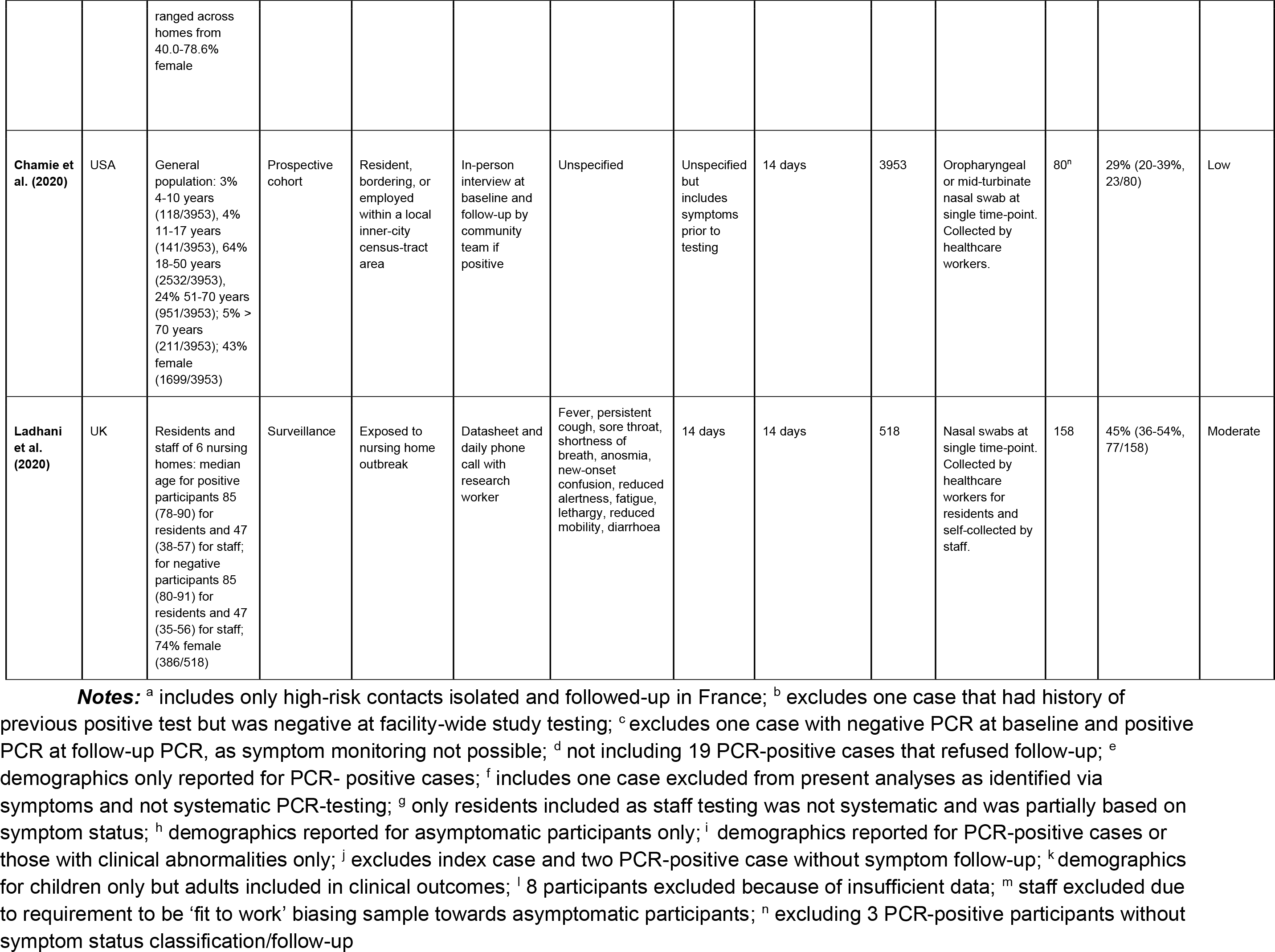
Descriptive Summary of Studies Included in Meta-Analysis.

**Figure 4.**
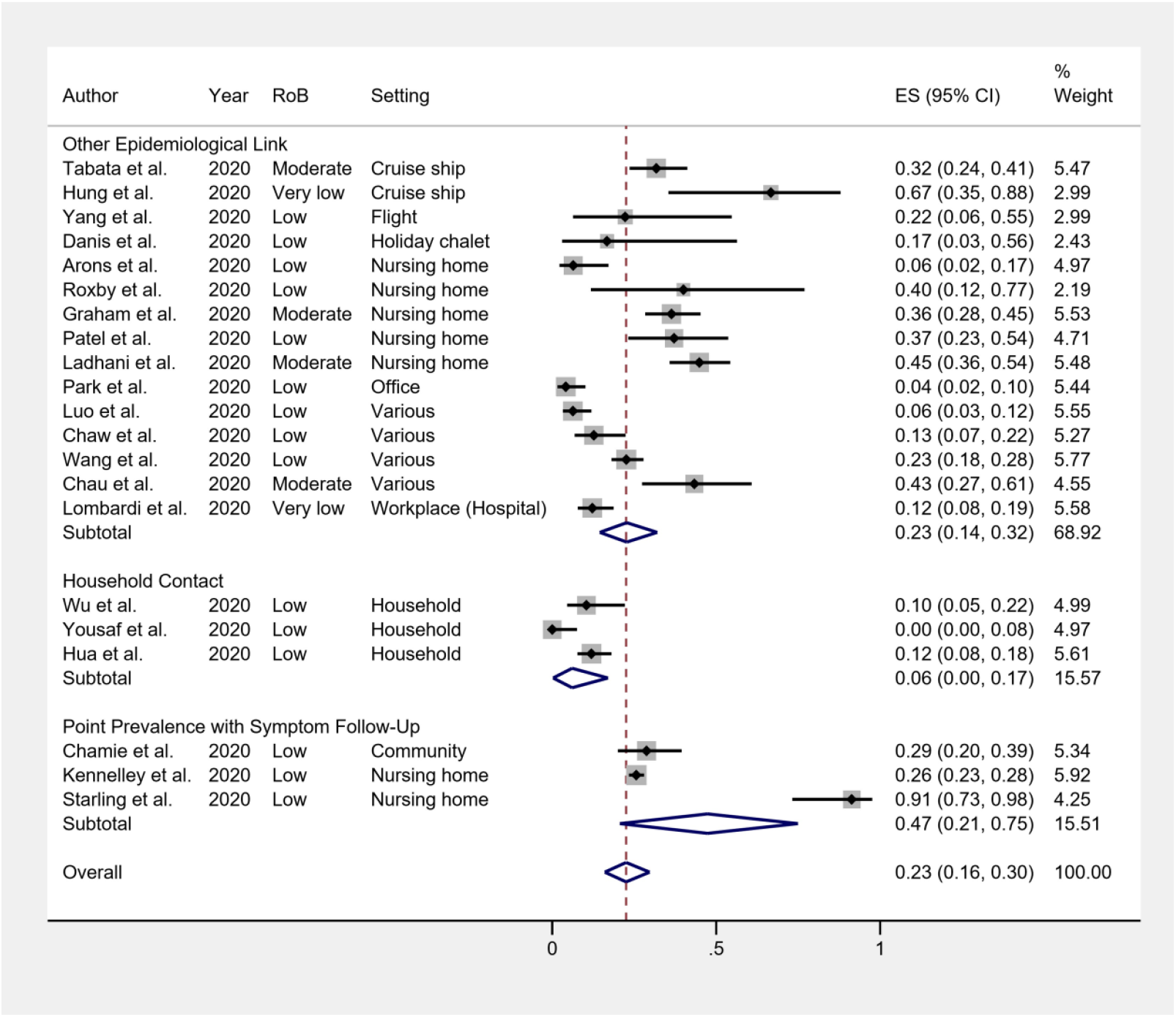
Meta-Analysis Results for COVID-19 Asymptomatic Proportion in Community Studies. ***Note:*** ES (effect size) = asymptomatic proportion; I^2 = heterogeneity; asymptomatic proportions are given in decimal form

The funnel plot (Figure 5) and Egger’s test did not indicate publication bias across studies included in the meta-analysis: *t*=0.23, *p*=0.82, 95% CI: −0.97, 1.20.

**Figure 5.**
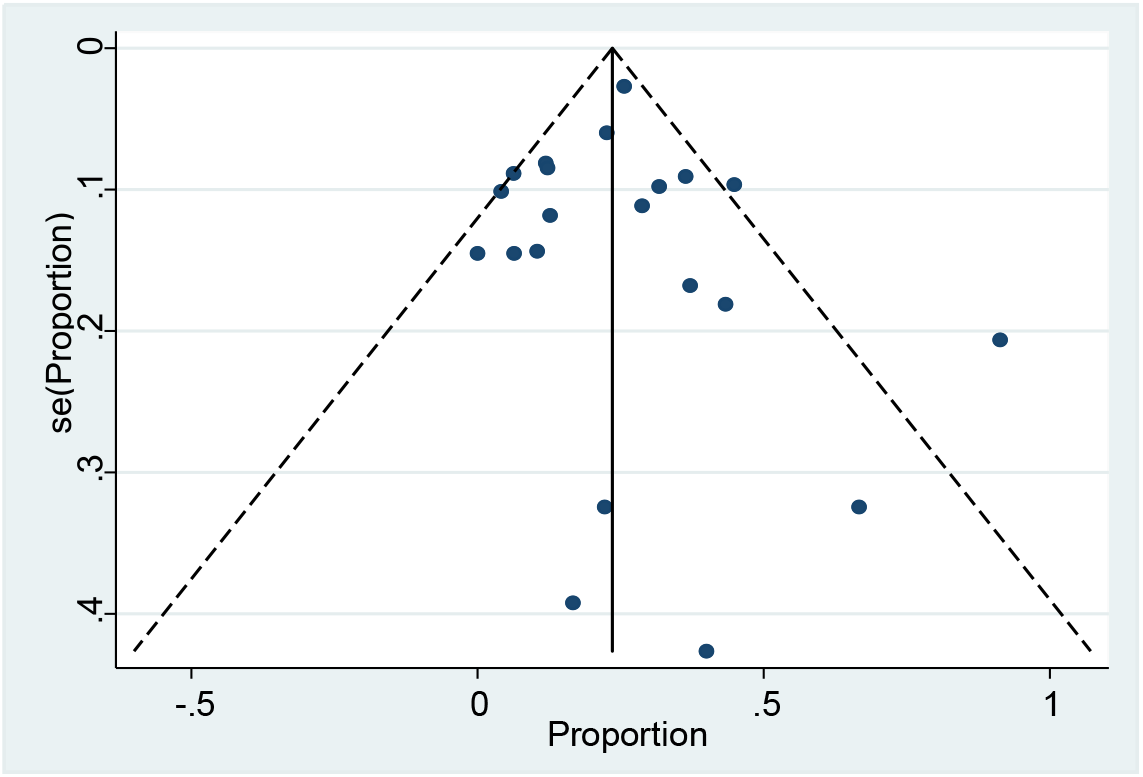
Funnel Plot for Meta-Analysis of COVID-19 Asymptomatic Proportion in Community Studies.

### Viral Load and Duration of Viral Shedding

Eight of the twenty-one included studies reported data regarding the CT values/viral load and/or duration of viral shedding for asymptomatic cases versus pre-symptomatic cases and/or those symptomatic from baseline. Differences in methodology and reporting precluded meta-analysis.

Five studies reported CT values and/or genome copy number by symptom status. One of these studies, Hung et al. (2020)^33^, found lower median baseline genome copy number in asymptomatic (3.86 log10 copies/mL) than symptomatic participants (7.62 log10 copies/mL). The remaining four studies all reported similar CT values for asymptomatic and symptomatic participants. Arons et al. (2020)^18^ reported similar baseline median cycle threshold values (CT) for asymptomatic (CT =25.5), pre-symptomatic (CT=23.1), and symptomatic (CT=24.5) cases. Infectious virus was isolated by viral culture from 33% (1/3) of available asymptomatic case specimens, 70.8% (17/24) of pre-symptomatic case specimens, and 65.0% (16/20) for symptomatic case specimens^18^. Chamie et al. (2020)^36^ also found that median CT values across samples were not significantly different between asymptomatic (CT=24, IQR: 19-26) and symptomatic individuals (CT=24, IQR: 19-25). Pre-symptomatic individuals appeared to have higher median CT values if seronegative and similar values if seropositive, but numerical detail was not reported overall for this group. Ladhani et al. (2020)^37^ also found no significant difference in baseline CT values between asymptomatic, pre-symptomatic, symptomatic, and post-symptomatic (i.e. reported symptoms in the two weeks prior to positive PCR result) participants; exact values were not provided. Chau et al. (2020)^21^ also reported similar baseline cycle threshold values for asymptomatic and symptomatic cases, though further numeric detail was not reported. When including all PCR results across follow-up for asymptomatic versus symptomatic cases (including negative PCR results), asymptomatic cases appeared to demonstrate lower CT values overall, which was proposed to indicate faster viral clearance^21^.

Direct investigation of duration of viral shedding was limited. Lombardi et al. (2020)^30^ found that median duration from positive test to first negative test was shorter in asymptomatic participants (22 days; IQR: 15–30) than symptomatic ones (29 days; IQR: 24–31), but the difference was not statistically significant. Danis et al. (2020)^20^ reported that the single asymptomatic case demonstrated the same viral load dynamics as one of the five symptomatic cases, with respective viral shedding periods of 7 and 6 days.

### Age of Symptomatic versus Asymptomatic Cases

Six studies^21,27,29,31,33,37^ reported information regarding the age of asymptomatic versus symptomatic cases. Variation in measurement and reporting precluded meta-analysis. Findings are reported in Table 3. Three studies indicated no significant difference in age between symptomatic and asymptomatic cases, while three studies suggested that asymptomatic cases tended to be younger than those with symptoms. Five studies were conducted in general population samples (contacts/potential contacts of confirmed cases or returning travellers), and one study was conducted in nursing home residents and staff with results stratified for these groups. Only one study^29^ reported a substantial child sub-sample (<14 years old), and found a higher asymptomatic proportion for infected children (23% *n*=10/43) than adults (7%, *n*=8/108).

**Table 3.** Reported Findings for Age of Asymptomatic versus Symptomatic Cases.

| Study | Sample | Findings |
| --- | --- | --- |
| Chau et al. (2020) | General public (contacts of confirmed case or returning travellers) | Median age of asymptomatic versus symptomatic participants: 30 (range 16-60) versus 27 (range 18-58) |
| Yang et al. (2020) | General public exposed to index case on flight | Median age of asymptomatic and symptomatic participants: 26 (IQR: 25.5-26.5) versus 33 (IQR: 29-45)<br>*note: very small asymptomatic sample ($n=2$) |
| Hua et al. (2020) | General public exposed to household cases or returning from high-risk areas | 23% of infected children ($\leq 14$ years, $n=10/43$) were asymptomatic versus 7% of infected adults ($n=8/108$), with children comprising 56% ($n=10/18$) of asymptomatic cases and adults 44% ($n=8/18$) |
| Tabata et al. (2020) | General public exposed to outbreak on cruise ship | Median age of asymptomatic versus symptomatic participants: 70 (IQR: 57-75) versus 68 (IQR: 56-74) |
| Hung et al. (2020) | General public exposed to outbreak on cruise ship | Median age of asymptomatic and symptomatic participants: 57 (IQR: 47-59) versus 68 (IQR: 59-68) |
| Ladhani et al. (2020) | Nursing home residents and staff | Median age of asymptomatic, post-symptomatic, pre-symptomatic, and symptomatic residents: 84 (IQR: 78-90); 88 (IQR: 85-91); 84 (IQR: 80-91); 87 (IQR: 80-91)<br>Median age of asymptomatic, post-symptomatic, pre-symptomatic, and symptomatic staff: 50 (IQR: 40-56); 54, (41-59); 38 (IQR 34-49); 40 (IQR: 26-55) |

## Discussion

Accurate estimates of the asymptomatic proportion of SARS-CoV-2 infections depend on appropriate study designs that systematically detect asymptomatic viral shedding and follow these cases up to differentiate truly asymptomatic infections from pre-clinical shedding. We calculated that an overall estimate of 23% of PCR-confirmed SARS-CoV-2 infections in community settings were asymptomatic, with a 95% confidence interval between 16%-30%. These findings do not support claims ^6,7,8^ of a very high asymptomatic proportion for PCR-confirmed infections (up to 80%) and highlight the importance of distinguishing between asymptomatic and pre-symptomatic cases. Heterogeneity in estimates of the asymptomatic proportion, however, was partly influenced by variation between testing contexts. Subgroup estimates range from 6% (95%CI 0-17%) for household contacts, increasing to 23% (95% CI 14-32%) for participants with other epidemiological links to case(s) or outbreaks, and the highest estimate of 47% (95% CI 21-74%) for point prevalence studies not directly linked to contact(s)/outbreaks.

These findings should be interpreted with caution in terms of the relationship between exposure and symptom status^38^. The assumption that household contacts of index cases may experience frequent and intense exposure with limited protection compared to other groups, and conversely that participants in non-outbreak studies may have more limited exposure, could not be empirically verified in the present review. Confidence intervals for subgroup asymptomatic proportions overlapped substantially, and data were limited for both the household contact and the point prevalence survey with symptom follow-up categories (both *n*=3 included studies). Furthermore, the estimate for point prevalence surveys was affected by one study^35^ with a very high asymptomatic proportion (91%); this estimate was likely influenced by the limited symptomatic case definition of new-onset cough or fever. Estimates for the other two studies were similar to the ‘other epidemiological link’ category (26% and 29%). Only one of the point prevalence studies with symptom follow-up^36^ was conducted in a general population sample. Furthermore, the ‘other epidemiological link’ category comprised a variety of study testing contexts, including studies that combined household contacts with participants with less intensive exposure, which likely contributed to substantial within-category heterogeneity. Despite these substantial limitations, further investigation is warranted into variability in the asymptomatic proportion across testing contexts as more data become available.

This effect of study context may partially account for differences between the overall estimate of the asymptomatic proportion in the current review and higher estimates from other studies. Notably, early population-based data collected from English households by the Office for National Statistics suggested that only 22% (95% CI 14-32%) of the 88 individuals who tested positive for COVID-19 thus far reported any symptoms, rising to 29% (95% CI 19-40%) of the 76 individuals tested repeatedly^8^. Similarly, 69% of another English community sample recruited regardless of symptom status reported no symptoms in the seven days up to their positive PCR result^39^. However, neither of these studies systematically followed-up cases regarding their symptoms across the course of infection, potentially overestimating the asymptomatic proportion and precluding inclusion in this review. Furthermore, findings were affected by the small sample size and consequently wide confidence intervals due to testing at a period of relatively low COVID-19 incidence in the population, as well as potential false positive PCR tests leading to an overestimate of asymptomatic cases. While some of these issues may have impacted studies included in the present review, the careful screening of study design and methodology done as part of this review was reflected in the overall very low or low risk of bias on assessed criteria for all but four included studies. An additional strength of our review is the systematic search of both peer-reviewed published literature and preprint studies which has enabled us to capture the most up to date estimates available.

Although this review identifies PCR-confirmed cases, PCR-confirmation and symptom-status alone cannot establish whether cases are infectious and, if so, the degree or duration of their infectiousness. Case reports, however, have indicated potential transmission of SARS-CoV-2 from some asymptomatic index cases ^1,2,9,21^. The balance of evidence regarding viral load in the present review indicates that asymptomatic cases had similar baseline or overall median viral loads to pre-symptomatic and symptomatic cases. Virological evidence suggests that infectious SARS-CoV-2 can be isolated by viral culture from samples with cycle threshold values up to 33, though the proportion of infectious virus decreases at higher cycle threshold values (i.e. lower viral load)^40^. While median baseline cycle threshold values for all symptom status groups (23.1-25.5) reported by Arons et al. (2020)^18^ fell well within this limit, infectious virus was isolated from only 33% of asymptomatic baseline samples, compared to 71% of presymptomatic and 65% of symptomatic samples. These findings should be interpreted with caution given the very small sample of asymptomatic specimens (*n*=3). Overall, clear reporting of cycle threshold values across follow-up by symptom status was lacking in included studies. This is an important area for further research given that the degree and duration of the infectious period for asymptomatic cases, as well as the overall proportion of virus-shedding cases that are asymptomatic, influence the contribution of asymptomatic cases to SARS-CoV-2 transmission at a population level.

Evidence regarding the duration of SARS-CoV-2 shedding by symptom status was very limited, with two studies suggesting no substantial difference in viral clearance times for asymptomatic and symptomatic cases. Duration of shedding varied widely between participants across all symptom status groups in included studies. The sample of asymptomatic cases in studies that reported duration of viral shedding also tended to be small, and the natural history of viral excretion by symptom status remains unclear. Further inquiry into the degree of preclinical shedding for pre-symptomatic cases, although not the focus of this review, is also warranted. The contribution of asymptomatic and pre-symptomatic cases to the overall spread of infection cannot be accurately inferred in the absence of high-quality evidence assessing the infectiousness of such cases^41^.

Evidence was also split regarding age and symptom status, with three studies indicating no difference in age between asymptomatic and symptomatic cases and three studies indicating that asymptomatic cases may tend to be younger than those with symptoms. Samples in the present study – both within the agerelated analysis and in the meta-analysis overall – tended to comprise primarily or exclusively of adults, and one study with a substantial child subsample^29^ found that a larger proportion of infected children were asymptomatic (23%) than adults (7%). Further comparison of the asymptomatic proportion for children and adults is required.

An important limitation of this review was the variability between symptomatic case definitions across included studies. Only eight of the twenty-one included studies^18,22-24,30,32,35,37^ described the full range of symptoms included within their symptomatic case definitions, while a further ten studies^19,20,23,25-29,33,34^ reported details of symptoms endorsed by participants but did not specify whether or which additional symptoms were assessed as part of their case definitions and three^17,31,36^ provided no detail. While a similar range of symptoms appear to have been monitored/endorsed across most included studies, it is possible that symptomatic case identification may have been affected by reporting bias and consequently that the true proportion of symptomatic cases was underestimated. Notably, Starling et al. (2020) ^35^ – the study with the highest reported asymptomatic proportion (91%) – used a very limited case definition of new-onset cough or fever. The reported proportion likely reflects individuals not meeting this case definition and excludes cases with other symptom profiles. This issue is particularly relevant given that unusual symptoms such as dysosmia/anosmia - only explicitly investigated by four studies^21,28,30,37^ - and dysgeusia/ageusia-only explicitly investigated by two studies^28,30^ - may be the primary or sole symptom for some COVID-19 cases ^42-44^. Demographic reporting across studies was also limited and it was not possible to stratify findings by further demographic characteristics. Estimates of the asymptomatic proportion may vary across population subgroups and this is a relevant area for future enquiry.

We included only studies with symptom-related follow-up to prevent symptom status misclassification. However, overestimation of the asymptomatic proportion may still occur in contact tracing studies initiated during established outbreaks, such as Graham et al. (2020)^24.^, if baseline symptomatic participants are classified as index cases and systematically excluded from the asymptomatic proportion. This review was also limited to estimating the asymptomatic proportion of virologically-confirmed infections. The asymptomatic proportion of infection varies depending on whether infections are identified using virological or serological methods^45^. PCR confirmation, which identifies infection with viral shedding, is informative for modelling transmission potential. However, review of the asymptomatic proportion of total infections based on emerging serological evidence – which identifies infections regardless of viral shedding – will be informative to understand how far SARS-CoV-2 has spread within populations and investigate evidence of immunity following asymptomatic infection^46^.

Overall, this review provides preliminary evidence that, when investigated using methodologically-appropriate studies, a substantial minority of SARS-CoV-2 infections with viral shedding are truly asymptomatic. These findings indicate that testing should not be exclusively limited to symptomatic individuals. Further research identifying distinguishing features (e.g. age) and testing contexts for truly asymptomatic cases, as well as their transmission potential, is recommended to inform testing programmes. These findings also highlight the importance of other public health measures, such as promoting social distancing and wearing face coverings in public places, regardless of symptom status.

## Data Availability

Data for this project have been made available at http://doi.org/10.5522/04/12344135

## Data Availability

Data for this project have been made available at http://doi.org/10.5522/04/12344135

## Data Availability

Data for this project have been made available at http://doi.org/10.5522/04/12344135

## Data Availability

Data for this project have been made available at http://doi.org/10.5522/04/12344135

## Funding

This work was supported by an MRC doctoral studentship (MR/N013867/1) to SB and a Wellcome Trust Clinical Research Career Development Fellowship to RWA (206602). The funders were not actively involved in the design, delivery, or analysis of this research. The views expressed in this publication are those of the authors and not necessarily of the MRC or the Wellcome Trust. AH is an NIHR Senior Investigator. The views expressed in this Article are those of the authors and not necessarily those of the NIHR, or the Department of Health and Social Care.

## Competing Interests

AH serves on the UK New and Emerging Respiratory Virus Threats Advisory Group. The other authors declare no competing interests.

## Data Availability Statement

University College London Research Data Repository: http://doi.org/10.5522/04/12344135 This project contains the following underlying data:

Asymptomatic meta-analysis V2.csv (data used to conduct meta-analysis of asymptomatic proportion) Data are available under the terms of the Creative Commons Attribution 4.0 International license (CC-BY 4.0).

## Notes

### Author Declarations

This research did not require ethical clearance as it comprised a rapid review and meta-analysis conducted using data reported in published/pre-printed articles only.

